# Preconception advice, care and services in the UK: who delivers what, where, how and to whom? A cross-sectional survey

**DOI:** 10.64898/2026.07.24.26358840

**Authors:** Danielle Schoenaker, Emma Hope Cassinelli, Oluwakemi Akagwu, Shivali Lakhani, Madeleine Benton, Lizzie Blundell, Sinead Brophy, Sinéad Currie, Jennifer Hall, Stephanie Hanley, Kate Maslin, Majel McGranahan, Cheryl McQuire, Judith Stephenson, Ruth Tunn, Laura McGowan, the UK Preconception Partnership

**Affiliations:** Lifecourse Epidemiology Centre, School of Human Development and Health, Faculty of Medicine, University of Southampton, Southampton, UK; NIHR Southampton Biomedical Research Centre, University of Southampton and University Hospital Southampton NHS Foundation Trust, Southampton, UK; School of Health Sciences, Faculty of Health and Medical Sciences, University of Surrey, Guildford, UK; School of Pharmacy, Applied Sciences and Public Health, Robert Gordon University, Aberdeen, UK; Department of Psychological Medicine, Institute of Psychiatry, Psychology & Neuroscience, King’s College London, London, UK; The National Perinatal Epidemiology Unit, Nuffield Department of Women’s and Reproductive Health, University of Oxford, UK; Population Health Data Science, Swansea University, Singleton Campus, Swansea, Wales, UK; Psychology, Faculty of Natural Sciences, University of Stirling, Stirling, UK; UCL EGA Institute for Women’s Health, London, UK; Department of Applied Health Sciences, College of Medicine and Health, University of Birmingham, Birmingham, UK; School of Nursing and Midwifery, University of Plymouth, Plymouth, UK and Department of Development and Regeneration, KU Leuven, Leuven, Belgium; Warwick Medical School, University of Warwick, Coventry, UK; Centre for Public Health, Bristol Medical School, University of Bristol, Bristol, UK; Centre for Public Health (Institute of Global Food Security), School of Medicine, Dentistry and Biomedical Science, Queen’s University Belfast, Belfast, UK

**Keywords:** preconception care, national health service

## Abstract

**Objective:** to describe the provision of preconception care across publicly funded or contracted health and social care settings in the UK.

**Design and setting:** online cross-sectional survey conducted October-December 2025.

**Population:** healthcare professionals delivering preconception care, recruited via professional organisations and networks.

**Methods:** quantitative data were analysed using descriptive statistics and qualitative free-text responses using inductive content analysis.

**Outcome measures:** preconception care content, target population, frequency of provision, funding and commissioning models, and approaches for reporting and monitoring.

**Results:** Eighty-seven healthcare professionals completed the survey. Most were women (89.3%), aged 41-60 (63.1%) and based in England (84.5%). Participants represented diverse roles, mainly obstetric/maternal-fetal specialists (23.0%), specialist nurses (16.0%), GPs and midwives (13.8% each). Preconception care primarily targeted women ≥20 years (98.9%), with fewer targeting men and adolescents. Care was usually embedded within relevant consultations (69.4%), particularly contraception, medication and health condition reviews, and often a one-off interaction (75.3%). Content focused on condition-related management/medication (68.6%), folic acid (66.3%), risky behaviours (smoking, alcohol, illicit drugs) (40.7%), diet (37.2%) and weight (36.0%). Services were mostly not formally commissioned (62.4%), lacked financial incentives (84.7%) and had no audit/service evaluation requirements (81.2%).

**Conclusions:** Preconception care in the UK is delivered by a wide range of healthcare professionals. Their engagement has improved considerably when compared with studies conducted over a decade ago, but preconception care remains fragmented, opportunistic and poorly supported by commissioning and system infrastructure. Strengthening integrated care pathways, funding mechanisms and use of standardised resources is essential to achieve consistent and equitable preconception care.

## Introduction

Addressing modifiable risk factors before pregnancy - such as poor glycaemic control, teratogenic medication use, suboptimal nutrition, smoking and alcohol consumption - can reduce the risk of adverse health outcomes, including congenital anomalies, pre-eclampsia and preterm birth.^1-6^ It also supports longer-term health for parents and children.^2,5,6^ Care and advice to optimise preconception health is ideally embedded across the health system and offered to all individuals of reproductive age as a substantial proportion of pregnancies are unplanned,^7^ and individuals may interact with a range of healthcare professionals throughout their reproductive years.^8^

Preconception care can be delivered by multiple professional groups working across the health system, including primary care, maternity, community and specialist services.^9^ In the UK, national policies and clinical guidelines recommend that preconception care should be integrated into routine clinical practice and delivered opportunistically across healthcare encounters, with the content tailored according to an individual’s health needs, reproductive intentions and risk factors.^10^ Core components include risk assessment, health promotion, optimisation of long-term conditions, medication review, and support for reproductive planning.^10^ These frameworks promote a multidisciplinary, life-course approach to improving health before pregnancy.^10,11^

Despite available policies and guidelines,^10^ there is limited understanding of how preconception care is implemented in day-to-day clinical practice. Existing research has largely focused on professional beliefs, attitudes and perceived barriers, rather than on how care is actually delivered within routine services.^12^ Studies from high-income countries, including England,^13,14^ Scotland,^15^ the Netherlands,^16,17^ and Australia^18^, suggest that preconception care is largely opportunistic, inconsistently structured, and primarily targeted at women with existing health conditions. However, these studies are restricted to primary care professionals, do not capture preconception care across the wider health system, and were mostly conducted at least 10 years ago.

A clear understanding of who provides preconception care, to whom, in what settings, and how it is commissioned, evaluated and supported is essential to inform service improvements and future research. This study aimed to describe the provision of preconception health advice and care across publicly funded or contracted health and social care settings in the UK.

## Methods

### Study design and participants

This study was a cross-sectional, self-completed online survey conducted between October and December 2025. Eligible participants were healthcare professionals and practitioners working in publicly funded (i.e. through the National Health Service [NHS]) or contracted health or social care settings in the UK who provide preconception health advice or care. Provision of preconception health advice or care was defined as: screening, risk assessment, education, counselling, treatment and/or advice provided to people of reproductive age, regardless of gender or pregnancy intention, with the aim of optimising health prior to a potential pregnancy.

Participants were recruited through professional organisations including the Primary Care Women’s Health Society, Royal College of Obstetricians and Gynaecologists (RCOG), College of Sexual and Reproductive Healthcare, Institute of Health Visiting, MacDonald Obstetric Medicine Society, among others (see acknowledgements). Professional organisations disseminated study invitations via eNewsletters, social media, or website postings. No targeted selection of individuals within organisations was undertaken. Invitations were distributed across all four UK nations. Additional participants were recruited via snowball sampling, with respondents encouraged to share the study within their professional networks.

We aimed to recruit n=75 participants from a range of clinical settings, including primary care, pharmacy, maternity services, sexual and reproductive health (SRH) services, health visiting, secondary care, and specialist services. This was an exploratory study that used a pragmatic sample size and sampling approach. We did not aim to obtain a representative sample of professional groups.

### Survey development and data collection

The survey was developed by a multidisciplinary team of researchers with expertise in preconception health and care (co-authors on this paper). It was piloted with 10 healthcare professionals across the four UK nations to assess clarity, relevance and acceptability.

Feedback on question wording and response options was incorporated prior to survey launch. The estimated completion time was 10-20 minutes.

The survey was administered via Qualtrics (**Supplementary File 1**) and included questions on:

- professional role and clinical setting
- content and target population of preconception care
- frequency of preconception care provision
- awareness, relevance and implementation of clinical guidelines and policies
- funding or commissioning models
- reporting and monitoring practices
- demographic characteristics

All closed multiple-choice questions were mandatory, and open-ended questions were optional to improve completion rates.

## Data analysis

Quantitative data were analysed using descriptive statistics to summarise participant characteristics and key aspects of preconception care provision. Results were described for the overall study population. Responses from ‘other’ categories were combined with existing response options (e.g. obstetrician reported as ‘other’ was combined with ‘obstetrician / gynaecologist’) or reported separately (e.g. ‘other: diabetes specialist nurse’). Qualitative free-text responses were analysed using inductive content analysis by coding responses into categories developed from the data. Coding of qualitative responses was done by EHC and verified by all co-authors. No further subgroup analyses were planned as part of this exploratory study due to the low pragmatic target sample size. Exploratory analyses by healthcare profession are presented in **Supplementary File 2** to inform potential future research.

This study is reported in line with Strengthening the Reporting of Observational Studies in Epidemiology (STROBE) guidance for cross-sectional studies.^19^ All quantitative summary statistics and coded qualitative data collected through the survey are reported in this paper. The anonymised original dataset is available online.^20^

## Results

### Participant characteristics

A total of 252 potential participants accessed the survey. Of these, 111 were exited via eligibility screening questions. Of 141 eligible participants, 94 provided consent and 87 completed the survey.

Most participants were aged 41-60 years (63.1%), predominantly women (89.3%) and of white ethnic background (82.1%) (**Table 1**). The majority were based in England (84.5%), with smaller representation from Northern Ireland (6.0%), Scotland (6.0%) and Wales (3.6%) (**Table S1**).

### Who provides preconception care and in what settings?

Participants represented a range of professional roles, most commonly obstetricians, obstetric physicians, gynaecologists and maternal-fetal-medicine specialists (23.0%), followed by specialist nurses (16.0%), general practitioners (GPs) (13.8%), midwives (13.8%) and SRH doctors (12.6%) (Table 1). Respondents worked across diverse settings, particularly maternity services (27.6%), hospital specialist services (18.4%), general practice (14.9%) and SRH services (13.8%). Over half (50.6%) had more than 16 years’ professional experience.

### Target populations for preconception care

Preconception care was primarily provided to women aged ≥20 years (98.9%), with lower engagement with men (31.0%) and non-binary or transgender adults (39.1%) (**Table 2**). Adolescents were less frequently included, particularly boys (11.5%) compared with girls (49.4%) and non-binary or transgender adolescents (26.4%).

Most respondents reported discussing preconception health with patients intending to conceive (70.9%), while fewer did so routinely with all or most individuals of reproductive age (46.5%) (Table 2).

When asked to describe their target population in more detail, open-ended responses (n=52 participants) indicated a strong focus on individuals with existing health conditions, particularly diabetes (n=11), epilepsy (n=5), fertility issues (n=5), alongside a combination of cardiovascular, renal, rheumatological, genetic and mental health conditions (n=20) (**Table S2**). Additional groups included those with previous adverse pregnancy outcomes, recurrent miscarriage, postnatal women, and socially vulnerable populations.

### Context, naming and frequency of preconception care provision

Preconception care was most often delivered when patients actively sought care (71.8%) or opportunistically within routine consultations (69.4%) (Table 2). Fewer participants provided preconception care through dedicated preconception clinics (28.2%). Among professionals who opportunistically embedded preconception care into relevant consultations, this was most frequently delivered within contraception (75.4%), health condition (47.4%) and medication reviews (45.6%).

Preconception care was predominantly delivered face-to-face (97.7%) and most reported providing this type of care to patients at least weekly (52.4%). It was typically a one-off interaction (75.3%), although 55.3% also reported integration into ongoing care (Table 2).

Among participants who responded to the question “How do you name and advertise your preconception care service?” (n=40), half of participants reported that preconception care was not delivered as a distinct service but embedded within routine care without a formal name or promotion (n=20) (**Table S3**). Where formalised, services were typically described as “preconception counselling clinics”, “pre-pregnancy clinics”, or embedded within maternal medicine pathways. Access was primarily via GP or specialist referral or word of mouth rather than public-facing promotion.

### Content of preconception care and use of patient-facing resources

Participant responses (n=86) indicated a broad but largely consistent focus on clinical risk assessment and health optimisation. The most reported components or topics for discussion were folic acid supplement use (n=57), behavioural factors including smoking, alcohol and illicit drug use (n=35) and diet and nutrition (n=32), and weight management (n=31) (**Figure 1, Table S4**).

**Figure 1.**
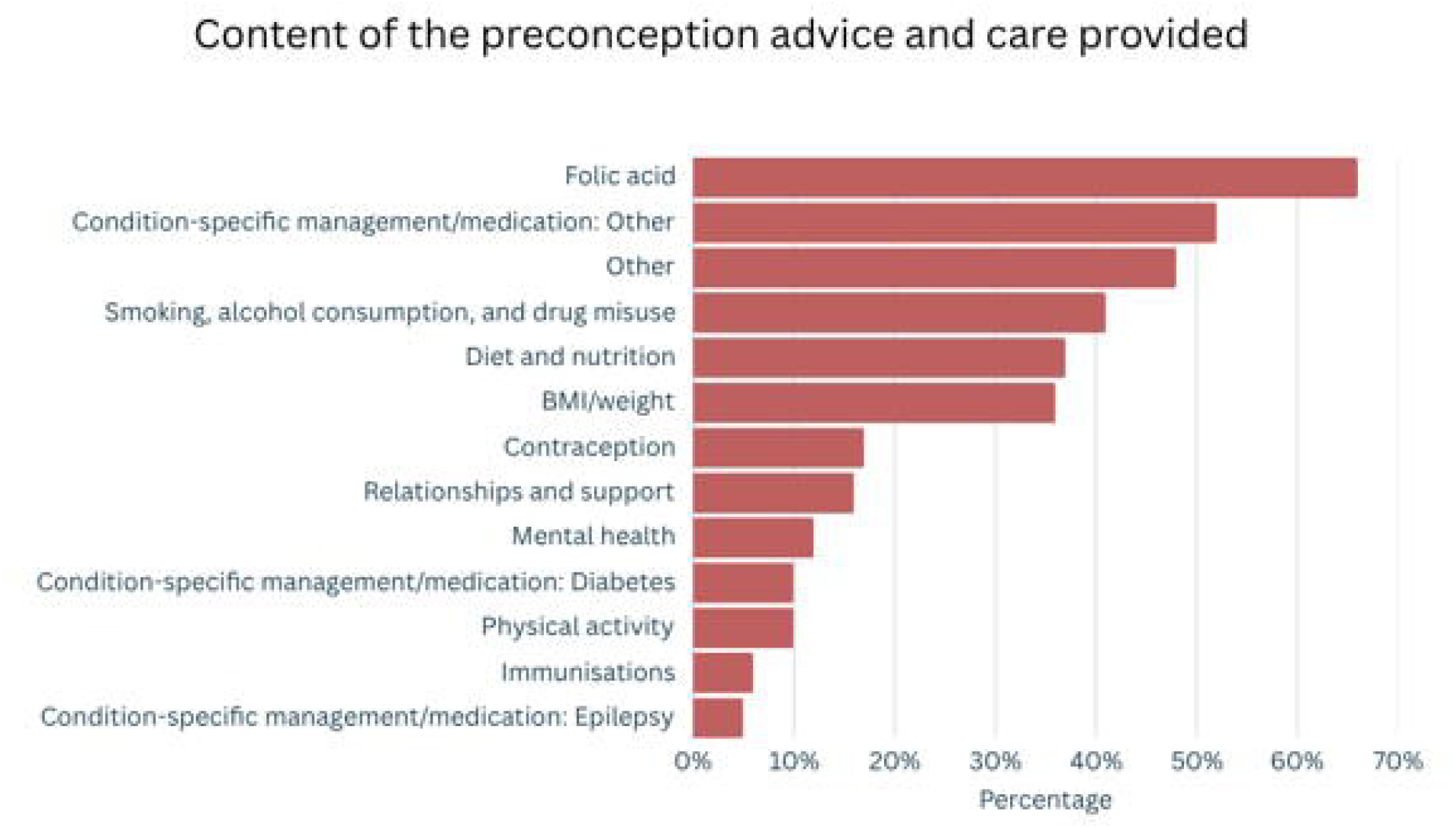
Content and topics discussed as part of preconception advice and care, N = 86

A substantial proportion of respondents described chronic condition-specific counselling, including for mental health (n=10), diabetes (n=9), epilepsy (n=4) and other/unspecified conditions and medications (n=45). This included medication review, avoidance of teratogenic drugs, and optimisation of disease control prior to pregnancy. Additional topics included contraception and pregnancy planning, physical activity, partner health and immunisations (Table S4).

Most participants used resources for patients, most commonly signposting to websites or apps (70.9%). Posters or leaflets (36.0%), checklists (15.1%) and social media (3.5%) were less frequently used, and 20.9% reported no use of patient-facing resources.

Commonly used digital resources, identified via open-ended responses (n=31), included the Tommy’s charity website (n=8) and national platforms such as the NHS website (n=6) (**Table S5**). Condition-specific resources were widely used for diabetes (Diabetes UK) and epilepsy (Epilepsy Action), including safety-focused guidance (e.g. medication risk and pregnancy prevention materials and websites) (**Table S6**).

### Guideline awareness and use

Most participants were aware of relevant preconception care guidelines or policies (72.9%). Among those, 19.6% did not use them in day-to-day clinical practice.

Based on open-ended responses (n=47), awareness was largely centred on National Institute for Health and Care Excellence (NICE) guidance (n=31), followed by RCOG guidance (n=13) (**Table S7**). Participants also reported a wide range of condition-specific guidelines (e.g. for diabetes, epilepsy, cardiovascular disease) (n=16), alongside a range of local policies and international guidance (n=19) (Table S7). A similar list was reported when participants were asked about those that they use in day-to-day clinical practice (**Table S8**).

### Funding, commissioning, evaluation and documentation

Preconception care was mostly not formally commissioned (62.4%), with smaller proportions funded through Integrated Care Boards (12.9%), NHS Boards (10.6%) or local authorities (5.9%) (**Table 3**). A further 15.3% of respondents were unsure of commissioning arrangements. Most respondents (84.7%) reported no financial or other incentives for provision.

Formal evaluation of preconception care provision was limited, with 61.2% reporting no evaluation (Table 3). Where evaluation occurred, it typically included audits, patient surveys and service-user feedback, often embedded within broader programmes such as maternal medicine services (**Table S9**). When specifically asked about reporting requirements for audit and/or service evaluation (“Are you required to report the preconception advice and care you provide for audit and/or service evaluation?”), 81.2% of participants indicated no formal requirements.

Two-thirds (68.2%) of respondents reported requirements to document care (“Are you required to document the preconception advice and care you provide as part of the clinical care process?”) (Table 3). This was most commonly done through electronic health records using free-text or structured templates (e.g. EMIS, SystemOne, EPIC, Ardens). Clinical letters to patients and GPs were also widely used (**Table S10**).

### Additional comments and reflections

Participants who left free-text responses at the end of the survey (n=17) frequently highlighted concerns about gaps in preconception care provision. Many described services as “*under-resourced*” and “*inconsistently prioritised*”, with “*reliance on voluntary or unfunded work*”. Preconception care was often described as occurring too late, typically once pregnancy had already been established. One participant noted it is “*extremely under-recognised*” and that there is “*still enormous ignorance around it*” (**Table S11**).

Inequities in access were also emphasised, with variation in service availability across regions and reliance on referral pathways, specialist networks or private care. This was described as *“a big equity issue as people in some parts of the UK are being offered preconception counselling and others are not”*.

Suggested improvements included structured tools or checklists to support delivery and earlier education, including introduction of preconception health into school curricula to improve awareness earlier in life.

## Discussion

### Main findings

This study provides the first system-wide overview of how preconception care is delivered across publicly funded and contracted health and social care settings in the UK. The findings show that preconception care is not structured, universally provided or consistently funded, but is instead delivered opportunistically by a range of professionals across primary care, maternity, specialist and SRH services. It most commonly occurs within existing consultations, particularly contraception reviews, medication management and long-term condition care. While largely informal, this suggests preconception care is embedded in multiple clinical touchpoints across the health system, providing a foundation for more coordinated, consistent and proactive approaches to be developed.

Our findings also show provision of preconception care is primarily focused on women, particularly those with existing health conditions, while engagement with men, adolescents and gender-diverse individuals was more limited. Formal clinics or services, where they exist, were inconsistently named, commissioned and promoted, and most care is provided without dedicated funding, financial incentives or service evaluation metrics. Despite this, clinicians across a range of roles reported integrating preconception advice into routine care and using clinical guidelines, templates, and national and local public-facing resources. Overall, preconception care in the UK remains fragmented and reactive, relying on individual clinician initiative rather than consistent service infrastructure.

### Interpretation considering other evidence

The finding that preconception care is delivered by a wide range of professionals across multiple settings aligns with growing literature and policy recognition that all health professionals who have contact with people of reproductive age have a role in providing preconception care.^10,12,21^ For example, since 2022, the Hatfield Vision for Women’s Health has recommended that comprehensive preconception care should be routinely offered during contraception consultations,^22^ reflecting evidence that women are receptive to discussing pregnancy intentions with SRH doctors.^23^ At the same time, shared responsibility across multiple professions without clearly defined roles may create ambiguity in ownership,^12,24^ contributing to the fragmented and inconsistent delivery observed in this study. Moreover, preconception care in our study was delivered predominantly by women (89.3%). This may partly reflect the composition of the NHS workforce,^25^ but it may also indicate that greater engagement from healthcare professionals, irrespective of their gender, is needed to normalise discussions about preconception health. Clearly defined, integrated care pathways, supported by appropriate education and training, are needed to equip healthcare professionals with knowledge, skills and confidence to deliver preconception care.

Consistent with previous research,^14,18^ preconception care was primarily targeted towards adult women, with some engagement of adolescent girls, but more limited inclusion of men and gender-diverse individuals. This contrasts with growing evidence on the importance of paternal preconception and life-course health,^2,5,6^ and the need for inclusive, tailored approaches for non-binary and transgender populations.^26,27^ Although most participants reported delivering care in response to pregnancy intention (70.9%), nearly half also provided it more routinely to all or most patients of reproductive age (46.5%). These findings may indicate an important shift in professional views and practice over the past decade. Earlier qualitative research conducted in England in 2011-2012 found that some healthcare professionals questioned whether preconception care formed part of their professional remit or service responsibilities. For example, one community SRH consultant stated that preconception care was “*not specifically part of our service specification*”, while another suggested that it was “*not a professional area*”.^28^ Similarly, a GP reported, “*I don’t do pre-pregnancy care - I tend to leave that for the antenatal appointments*”.^28^ Previous research in England also suggested that preconception advice was typically offered to women actively trying to conceive and rarely raised with men unless subfertility was a concern.^14^ In contrast, participants in the present study frequently described incorporating preconception discussions into routine consultations, which is important given low rates of pregnancy preparation, and supports a broader, life-course approach that normalises preconception health across the population.^11,29,30^

The strong focus on individuals with pre-existing conditions and medication use is consistent with clinical guidelines and reflects a more established role for specialist and secondary care services, in particular for conditions such as diabetes and epilepsy.^10,14,21,31^ The substantial representation of professionals working in maternity services in our study also highlights an important opportunity to support interpregnancy care. However, concentrating provision on these specific groups risks excluding individuals not in regular contact with healthcare and may reinforce disparities in access. This highlights the need for community, educational and population-level/public health strategies alongside healthcare-based interventions.^11,29,32^

The content of preconception care reported in our study - particularly folic acid supplement use, behavioural risk factors, diet and weight management - reflects clinical guidance and priorities identified in the literature.^10,21,33^ However, variation in coverage suggests that even core components are not consistently addressed. This aligns with previous studies,^14,17,18^ and reflects wider evidence of gaps in knowledge among health professionals.^12,14,15,18,24,34^ Notably, inconsistent and inadequate attention to folic acid and weight management contrast with worsening population trends in supplement uptake and maternal obesity.^35-37^ Broader topics, such as mental health and partner support, were mentioned by some participants, consistent with increasing recognition of their importance in both healthcare professionals and public perspectives.^2,33,38^

The use of patient-facing resources, particularly signposting to websites, may reflect wider trends towards digital information provision.^39,40^ However, one in five participants in our study reported not using such resources, consistent with previous findings of limited availability or use of patient materials.^12,18^ The reliance on a small number of trusted national and charitable resources suggests that while clinicians are actively seeking tools to support care, there is scope for improved availability and awareness of evidence-based resources. Contrary, awareness of clinical guidelines was relatively high (72.9% in our study), which may represent an improvement compared with earlier studies. For example, 53% of Australian GPs reported they were aware of preconception care guidelines in 2019,^18^ while GPs in an interview study conducted in England in 2016 were “*not familiar with preconception guidelines*”.^14^ However, one in five participants in our study did not use guidelines in day-to-day clinical practice (19.6%). Although it is unclear if this means these participants did not access guidelines, or if they did not deliver care in line with guidelines, it may reflect previous findings that implementation may be constrained by time pressures and lack of system-level support.^11,12,32^

Finally, the limited commissioning, funding and evaluation of preconception care reinforces concerns that, despite scientific evidence of its importance and the availability of clinical guidelines, preconception care is not adequately supported at a system level.^14,24,32,34^ Although preconception care is recognised within the service specification for Women’s Health Hubs in England,^41^ its integration into routine care appears inconsistent, reflecting the absence of dedicated commissioning and accountability mechanisms. The widespread use of electronic documentation systems suggests there is an opportunity to strengthen monitoring and evaluation,^13,38,42-45^ for example through quality improvement frameworks or incentivisation mechanisms, to support more consistent and equitable provision.

### Strengths and limitations

A key strength of this study is its system-wide perspective, capturing preconception care across multiple professional groups, settings and UK nations. The survey design included multiple-choice in combination with free-text responses to better understand preconception care target populations, content, resource use, evaluation and reporting. The inclusion of diverse clinical roles provides a broad overview of current service provision, and piloting of the survey improved clarity and reduced measurement error.

However, several limitations should be considered. The study sample is unlikely to be representative of all UK healthcare professionals delivering preconception care. Although recruitment across multiple organisations aimed to capture diverse participants, this may also have introduced selection bias, particularly towards clinicians working in primary care, obstetrics and SRH services. Participation was voluntary and may be subject to self-selection bias, with likely over-representation of individuals with a special interest in preconception care. Some participants reported uncertainty about the definition of preconception care, which may have limited participation among professionals who provide relevant advice but do not explicitly identify it as such. In addition, data were self-reported and therefore subject to recall and social desirability bias. Findings should therefore be interpreted as exploratory and descriptive rather than as definitive estimates of national provision. Given the likely over-representation of clinicians engaged with preconception care, these findings may present a more favourable picture of current practice than exists across the wider healthcare system.

## Conclusion

This study demonstrates that preconception care in the UK is delivered by a wide range of healthcare professionals but remains inconsistently provided and insufficiently supported by commissioning and system infrastructure. Although there is clear professional commitment, delivery is largely concentrated within long-term condition management, medication review and contraception-related consultations, leaving gaps for those not accessing care through these pathways.

Importantly, the findings suggest that professional engagement with preconception care has changed considerably since 2010, potentially reflecting growing policy and professional recognition of its importance. However, the findings highlight a fundamental mismatch between clinical activity and system-level support, with limited commissioning, incentives and formal service structures. Addressing this gap will be essential to shift responsibility for preconception care from reliance on individual patient presentations and opportunistic clinical encounters towards a more systematic, proactive and integrated approach within routine healthcare systems. Establishing commissioning frameworks, strengthening service integration, developing standardised care pathways and aligning trusted resources could support more consistent, equitable and life-course delivery of preconception care across the UK.

## Data Availability

All data produced are available online at https://doi.org/10.5258/SOTON/D3968.

## Acknowledgements

Acknowledgements: The authors would like to thank all healthcare professionals and practitioners who took part in the study, and organisations and individuals who supported participant recruitment by sharing information about the study, including the UK Preconception Partnership, Scottish Preconception Health Research Network, Scottish Government Preconception health and care collaborative, Healthier Pregnancies Better lives, Primary Care Women’s Health Society, WiseGP, General Practitioners Championing Perinatal Care, Society for Academic Primary Care, Oxford Primary Care network, Community pharmacy Scotland, Royal College of Midwives, College of Sexual and Reproductive Healthcare, Institute of Health Visiting, Royal College of Obstetricians & Gynaecologists, MacDonald Obstetric Medicine Society, British Dietetic Association, UK Association for the Study of Obesity, Diabetes in Pregnancy Working Group, Active Pregnancy Foundation, Epilepsy Nurses Association, Epilepsy Action, Epilepsy Research Institute UK.

## Contribution to authorship

DS and LMG conceived the idea. All authors contributed to survey design and participant recruitment. EHC, OA and SL contributed to survey set up in Qualtrics. EHC analysed data and created tables and figures. DS wrote the first draft of the manuscript. All co-authors contributed to reviewing and editing the manuscript.

## Ethics approval

This study has been approved by the University of Southampton Faculty of Medicine Ethics Committee (ERGO 104614).

## Table and figure caption list

Table 1. Participant socio-demographic characteristics, N = 87^1^

Table 2. Provision of preconception care: target population and service characteristics, N = 87^1^

Table 3. Funding, commissioning, evaluation and documentation of preconception care, N = 85^1^

