## Supplementary material for "Preconception advice, care and services in the UK: who delivers what, where, how and to whom? A cross-sectional survey": Tables

**Table 1**. Participant socio-demographic characteristics, N = 87^1^

| **Characteristic** | **n (%)** |
| --- | --- |
| **Age group** |  |
| 21-30 years | 12 (14.3) |
| 31-40 years | 14 (16.7) |
| 41-50 years | 25 (29.8) |
| 51-60 years | 28 (33.3) |
| 61-70 years | 5 (6.0) |
| **Gender** |  |
| Woman | 75 (89.3) |
| Man | 7 (8.3) |
| Prefer not to answer | 2 (2.4) |
| **Ethnic background** |  |
| White | 69 (82.1) |
| Asian/Asian British | 6 (7.1) |
| Indian | 6 (7.1) |
| Any other ethnic group^2^ | 3 (3.6) |
| **Work location** |  |
| England | 71 (84.5) |
| Northern Ireland | 5 (6.0) |
| Scotland | 5 (6.0) |
| Wales | 3 (3.6) |
| **Main profession** |  |
| Obstetrician, gynaecologist obstetric physician, maternal-fetal-medicine specialist | 20 (23.0) |
| General practitioner (GP) | 12 (13.8) |
| Midwife | 12 (13.8) |
| Sexual and reproductive health doctor | 11 (12.6) |
| Community or practice pharmacist | 5 (5.7) |
| Health visitor | 5 (5.7) |
| Specialist nurse: Diabetes | 5 (5.7) |
| Specialist nurse: Epilepsy | 7 (8.0) |
| Specialist nurse: Family Nurse Partnership | 2 (2.3) |
| Other; including psychiatrist, neurologist, cardiologist, geneticist | 8 (9.2) |
| **Years in main profession** |  |
| Less than 5 years | 19 (21.8) |
| 5-10 years | 4 (4.6) |
| 11-15 years | 20 (23.0) |
| 16+ years | 44 (50.6) |
| **Professional setting** |  |
| Maternity services (antenatal/birth/postnatal) | 24 (27.6) |
| Hospital specialist service | 16 (18.4) |
| General practice | 13 (14.9) |
| Sexual and reproductive health service | 12 (13.8) |
| Health visiting | 6 (6.9) |
| Community pharmacy | 5 (5.7) |
| Other; including mental health service, early pregnancy unit, fertility clinic, pre-pregnancy clinic, community NHS trust, Family Nurse Partnership | 11 (12.6) |

^1^ Sample size ranges from 84-87 due to missing values.

^2^ Any other ethnic group: includes responses Any other ethnic group, Bangladeshi, and Mixed/multiple ethnic groups.

### Table 2. Provision of preconception care: target population and service characteristics, N = 87^1^

| **Characteristic** | **n (%)** |
| --- | --- |
| **Target population: age and gender^2^** |  |
| Adolescent girls (10-19 years) | 43 (49.4) |
| Adolescent boys (10-19 years) | 10 (11.5) |
| Adolescent non-binary or transgender people (10-19 years) | 23 (26.4) |
| Women (20+ years) | 86 (98.9) |
| Men (20+ years) | 27 (31.0) |
| Non-binary or transgender people (20+ years) | 34 (39.1) |
| **Target population: pregnancy intention & other characteristics^2^** |  |
| Patients who are intending to conceive | 61 (70.9) |
| All / most patients of reproductive age | 40 (46.5) |
| Other: Patients with medical conditions / medication use | 4 (4.7) |
| Other: Postnatal women | 3 (3.5) |
| Other: Pregnant women | 2 (2.3) |
| **Reaching the target population^2^** |  |
| Patients/people actively seek advice and care | 61 (71.8) |
| Care and advice proactively embedded into relevant consultations | 59 (69.4) |
| Patients/people are referred through other healthcare professionals and services | 40 (47.1) |
| Specific preconception care service/clinic | 24 (28.2) |
| **If care and advice proactively embedded: relevant consultations^2,3^** |  |
| Contraception | 43 (75.4) |
| Health condition review | 27 (47.4) |
| Medication review | 26 (45.6) |
| Fertility | 25 (43.9) |
| Sexual health | 25 (43.9) |
| Routine health check | 23 (40.4) |
| Postnatal check | 21 (36.8) |
| Cervical smear | 12 (21.1) |
| Other, including antenatal care, healthy child programme, health visiting contacts, any / regular appointments, newly registered patient review, HIV appointment | 12 (21.1) |
| **Mode of care provision^2^** |  |
| Face-to-face consultation | 84 (97.7) |
| Online or phone consultation | 60 (69.8) |
| Promotion of campaigns/information | 18 (20.9) |
| SMS or email | 15 (17.4) |
| **Frequency of care provision** |  |
| Daily | 8 (9.4) |
| Weekly | 43 (50.6) |
| Monthly | 20 (23.5) |
| Less than monthly | 9 (10.6) |
| Other | 5 (5.9) |
| **Frequency of patient contact as part of preconception advice and care^2^** |  |
| One off consultation / conversation | 64 (75.3) |
| As part of ongoing care | 47 (55.3) |
| Other: Follow up if required | 2 (2.4) |
| Other: Varies or unclear | 5 (5.9) |
| **Years provided preconception advice and care as healthcare professional** |  |
| Less than 5 years | 35 (41.2) |
| 5-10 years | 9 (10.6) |
| 11-15 years | 12 (14.1) |
| 16+ years | 29 (34.1) |
| **Years current service provided preconception advice and care** |  |
| Less than 5 years | 9 (10.6) |
| 5-10 years | 8 (9.4) |
| 11-15 years | 10 (11.8) |
| 16+ years | 39 (45.9) |
| Don't know | 19 (22.4) |

^1^ Sample size ranges from 85-87 due to missing values.

^2^ Multiple answer options possible.

^3^ Only asked if response to question ‘How do you reach the target population?’ was ‘I proactively embed preconception advice and care into relevant consultations’.

### Table 3. Funding, commissioning, evaluation and documentation of preconception care, N = 85

| **Characteristic** | **n (%)** |
| --- | --- |
| **Commissioning of preconception advice and care or service^1^** |  |
| Not formally commissioned | 53 (62.4) |
| Integrated Care Board | 11 (12.9) |
| NHS Board | 9 (10.6) |
| Local authority / local council | 5 (5.9) |
| Don't know | 13 (15.3) |
| Other: Related services, including Fetal Medicine Unit consultation, postnatal contraceptive consultation, regular out-patient clinic consultation, perinatal mental health consultation, Maternal Medicine Network | 7 (8.2) |
| **Incentives in place (financial or other)** |  |
| No | 72 (84.7) |
| Yes | 3 (3.5) |
| Don't know | 10 (11.8) |
| **Evaluation of preconception advice and care or service** |  |
| No | 52 (61.2) |
| Yes | 18 (21.2) |
| Don't know | 15 (17.6) |
| **Audit and/or service evaluation required** |  |
| No | 69 (81.2) |
| Yes | 16 (18.8) |
| **Reporting requirement in place** |  |
| No | 27 (31.8) |
| Yes | 58 (68.2) |
| **If reporting requirement in place: method of documentation^1,2^** |  |
| Digital written notes | 53 (91.4) |
| Digital preconception care template | 9 (15.5) |
| Other: Letter to patients | 8 (13.8) |
| Other: Letter for referrals | 3 (5.2) |
| Other: Handwritten notes | 2 (3.4) |

^1^ Multiple answer options possible.

^2^ Only asked if response to question ‘Are you required to document the preconception advice and care you provide as part of the clinical care process?’ was ‘Yes’.
