## Supporting Information for "Preconception advice, care and services in the UK: who delivers what, where, how and to whom? A cross-sectional survey"

#### Table S1. Work location of study participants, N=84

| **Location** | **n (%)** |
| --- | --- |
| England - London region | 13 (15.5) |
| England - East Midlands region | 4 (4.8) |
| England - East of England region | 4 (4.8) |
| England - North East region | 3 (3.6) |
| England - North West region | 15 (17.9) |
| England - South East region | 10 (11.9) |
| England - South West region | 9 (10.7) |
| England - West Midlands region | 4 (4.8 |
| England - Yorkshire and the Humber region | 9 (10.7) |
| Northern Ireland - Belfast Health and Social Care Trust | 1 (1.2) |
| Northern Ireland - Northern Health and Social Care Trust | 1 (1.2) |
| Northern Ireland - Southern Health and Social Care Trust | 3 (3.6) |
| Scotland - NHS Grampian | 1 (1.2) |
| Scotland - NHS Greater Glasgow & Clyde | 1 (1.2) |
| Scotland - NHS Lanarkshire | 1 (1.2) |
| Scotland - NHS Lothian | 2 (2.4) |
| Wales - Betsi Cadwaladr University Health Board | 1 (1.2) |
| Wales - Cardiff and Vale University Health Board | 1 (1.2) |
| Wales - Hywel Dda University Health Board | 1 (1.2) |

#### Table S2. Further descriptions of preconception care and advice target populations, N=52

| **Category** | **Count** | **Responses** |
| --- | --- | --- |
| Diabetes | 11 | - Diabetes - Diabetes - Diabetes - Diabetes of any type in any pregnancy - Many different people with various conditions, diabetes […] - People with diabetes - Type 2 diabetes - Women with a diagnoses of diabetes of reproductive age - diabetes […] - diabetes preconception advice - Cystic fibrosis, diabetes |
| Epilepsy | 5 | - Epilepsy - Epilepsy, transition from child services, including people experiencing epilepsy and co-morbidity e.g. depression and anxiety disorders, dissociative seizures (sometimes referred to as functional/non-epileptic attacks) - Patients with epilepsy - People with Epilepsy - […] but I would focus on people of child bearing age. (epilepsy) |
| Other health conditions | 20 | - […] I tend to get referrals mainly for medical conditions […] - Cardiology patients so ACHD, ICC and acquired plus women who have been under obs cardiology clinic who have delivered - Cystic fibrosis - […] complex health patients - […] patients with complex medical conditions (haematology, rheumatology etc) - […] With women it is most likely to arise if prescribing specific medicines […] - […] We are also a regional referral centre for patients with complex medical problems wishing TOP (usually for social reasons) […] - I see women for preconception care with underlying renal disease before they intend to get pregnant and also when they have made the decision of intention to become pregnant. I also see women with Rheumatological disease in the same position - I work as an O&G trainee with an interest in maternal medicine. I work in a district hospital and and we are referred patients for preconception counselling with a range of medical conditions - Many different people with various conditions, diabetes, mental health - Medical conditions -cardiac, rheumatology, IBD, oncology - Patients with personal or family history of a genetic condition, as well as patients who would like to know whether a condition in the family has a genetic basis - Potentially any pre-existing medical problem or previous medical complication in pregnancy. Most common in my practice are cardiac disease, liver disease, cystic fibrosis, previous pre-eclampsia, which align with the antenatal clinics I provide. - Renal, Liver, Haematology, Rheumatology and Maternal medicine. We do not see women with Diabetes or Cardiac conditions - Those using secondary mental health services (including drug and alcohol services) - We discuss planning a family with all our patients (16+) if appropriate (we have a large population of people with an intellectual disability) […] - […] pcos medication inc glp1s - maternal medical or previous fetal conditions - women with medical disorders pre existing - women with medical problems (not surgical/gynaecological) |
| Pregnancy planning | 3 | - AFAB intending to conceive of any age - Considerations for future pregnancy planning - […] an opportunity to discuss the benefits of LARC and the need to access pre-conception care prior to embarking on a planned pregnancy |
| Fertility | 5 | - […] I do discuss PCC with patients referred to my Gynae OP for fertility issues as well […] - I do fertility clinic and a recurrent msicarriage clinic - I probably only discuss it with men if it specifically arises, such as in the context of subfertility. With women it is most likely to arise if prescribing specific medicines, etc, and asking about pregnancy intentions. Few people seem to consult for this, except in the context of subfertility - In my Endocrinology clinic I see a lot of patients who are struggling to conceive, male and female. I also discuss fertility plans with male and female patients of appropriate age […] - […] people in gyne/fertiltiy centre |
| General population | 3 | - Full General Practice population range - General - General childbearing population […] |
| Other | 16 | - 16-55 years - I work for an Adult service or patients 16 years plus - C Sections - Have not advised to people with specific health conditions - I run the termination of pregnancy service - I see women/couples before and after they have pregnancy so both pre and interconception consultations. The postnatal conversations usually follow an adverse pregnancy outcome at my hospital eg late miscarriage, preterm birth, fetal growth restriction. I receive preconception counselling referrals from GPs and hospital consultants to see women who have had an adverse pregnancy outcome at another hospital. I like to see both partners as the health message to men is important as well as to women. - […] In my role as an obstetric physician women are referred to me for pre-conception counselling and I discuss contraception with my pregnant patients prior to delivery. - Listening Service post natal debriefing - Mostly around LARC fittings or removals - Rainbow debriefs, GP referrals - Women who have just had a child - at the moment my contact is primiarly with either couples attending the early pregnancy unit, the recurrent miscarriage service or general antenatal care - first time teenage mothers, targeting post birth contraception and sexual health, we aim to delay/space subsequent pregnancy - homeless women - pregnant people […] - teenage pregnancy |

#### Table S3. Naming and advertisement of preconception care services, N=40

| **Category** | **Count** | **Responses** |
| --- | --- | --- |
| None, ad hoc or as part of care | 20 | - We don't advertise it - we don't, it's just general practice - don't advertise - we don't but we are an MMC - we dont - We don’t have a specific service - We don't advertise, we make patients aware as part of their regular consultations […] - Its not a dedicated service - […] this is not actually applicable because the service doesn’t provide preconception health advice - I do the other pharmacists don’t necessarily - we don't - we don't - Ad hoc - Not a specific+2 service, people organically present at pharmacy seeking support - Preconception Conversations - I am the women's health lead at the GP practice so patients often seek me out, I do the PN checks so will discuss it in this consultation. - Maternal health and well being discussion in pregnancy and ongoing in postnatal period - no specfic servcie part of my range of services as GP - As part on ongoing care. Listed in service leaflet and on the service website - Always just ad hoc - There is no set clinic. Currently the service runs as hoc when we can slot patients in during MDT/admin time |
| Clinics | 11 | - It is part of the UCLH Preterm Birth clinical service. https://www.uclh.nhs.uk/our-services/find-service/womens-health-1/maternity-services/your-pregnancy/preterm-birth-clinic - Debrief clinic - PRC PREGNANCY RISK CLINIC - Preconception counselling clinic - The maternal medicine centre pre-con clinic is advertised through our maternal medicine network and is included in our regional guidelines - We have a dedicated pre pregnancy counselling services at Nottingham that is jointly run by maternal medicine consultants and obstetric physicians in the 3rd Monday of every month. We have a dedicated admin team and inbox where we receive the referrals for PPC from primary and secondary care - PPCC pre pregnancy counselling clinic. […] - pregnancy risk clinic (a generic name for all women we see for additional counselling and includes post pregnancy loss and pre term birth as well) - "Formal preconception clinic advertised to primary care services and wider maternal medicine network - Women's Health / Pre Pregnancy clinic - Preconception counselling clinic |
| Referrals or word of mouth | 8 | - Patients can be referred through the maternal medicine centre referral form (online form) which can be accessed by GP, specialist physicians, specialist nurses,subfertility clinics - a referral service from the gp - referral from physicians in the hospital - […] Word of mouth via other Doctors and specialists - word of mouth - GP`s who have trained in the unit" - Mainly by word of mouth and colleagues who are aware refer onto my service. Also, offer self referral for patients who have previously used the service. - […] patients referred in via GP or other Consultants |
| Other | 8 | - I have worked locally with GPs, cardiology colleagues in my trust and neighbouring DGHs, midwives, obs team and the regional genetics team - via Trust/ PHA - we are considering rebranding - and doing all the pre conception cases in one session and named that - Specialist advice for neurology patients available to all practitioners in primary / secondary case care aware but no specific name / advertisement of this to my knowledge" - Preconception Care - Mainly leaflets - dont know - It is part of our holistic care, our consultants know that we have access to joint obs/epilepsy clinic and that we can ask the consultant obstetricians to do joint appointments as well if people have specific concerns |

#### Table S4. Content of preconception advice and care, N=86

| **Category** | **Count** | **Responses** |
| --- | --- | --- |
| Condition-specific management/medication: Diabetes | 9 | - […] diabetes and other meds review with endocrine team […] HbA1c checks and targets, considering the use of time in pregnancy range target glucose levels (for those with pre-existing diabetes) […] asking endocrinologists for pregnancy suitable insulin pump therapy - Diabetes preconception advice following Nice guidance […] stopping in appropriate medications, starting insulin - Diabetes specific facts related pregnancy - […] importance of getting optimal diabetes control, risk of diabetes on pregnancy, how pregnancy can affect diabetes, support with improving diabetes […] - […] glucose control, reasons for glucose targets for pregnancy, what to expect in a pregnancy that includes diabetes - […] insulin pump therapy. Glycaemic control - […] Also include it as part of medical history assessment ref to Diabetes […] - […] the importance of a good HbA1c first. - […] hba1c\ blood glucose targets […] |
| Condition-specific management/medication: Epilepsy | 4 | - ASM use in pregnancy and the risks of seizures if medications are stopped suddenly, use of high dose folic acid for women with epilepsy, the teratagenic risk of ASM against background risk, including the neurodevelopmental risk - Epilepsy review of diagnosis, medication and seizure control. Folic acid advice, risk assessment and prevention. Contraception advice and information about specific epilepsy drug interactions. - Also include it as part of medical history assessment ref to […] epilepsy […] - […] Discuss their epilepsy control, teretogenicity and options for their treatment regime (eg stay the same, or make changes to reduce risk to the baby) […] |
| Condition-specific management/medication: Other/unspecified | 45 | - […] medications […] if medical condition ideally well controlled - Current status of medical condition and implications for pregnancy. Effect of medical problem on pregnancy / associated risks. Medication review and suitability for pregnancy, alternative treatment options if current treatment unsuitable. Plan for management of pregnancy / management of medical problem in pregnancy, including birth options. […] - […] other medical conditions or medications - […] Medicine review to ensure safe to take in pregnancy […] - […] medicines review - […] avoiding teratogenic medication - […] medical conditions […] - […] any chronic medical conditions that may need review - […] medication review […] - […] correct medication […] - […] medication […] - […] medication […] - […] medication review, medical condition review […] - […] potential impact of disease on fertility/pregnancy and impact that pregnancy may have on disease control, medication review and risk of MCM if relevant […] - […] safe medication review […] - […] ensuring regular meds are pregnancy safe - […] medications in use […] - […] How pregnancy might affect the medical condition, how the condition might affect pregnancy, medications, genetics, fertility, pregnancy pathway, any investigations/optimisation that it would be beneficial to have prior to pregnancy - I discuss impact of preg on cardiac condition, impact of cardiac condition on preg, ensure up to date assessment, advise on timing of pregnancy, advise on best way to obtain optimum health prior to conception eg lifestyle etc and that includes folic acid, I also discuss the impact of the pregnancy eg visits, medical versus midwife led care, any delivery implications, discuss impact of pre-term delivery if appropriate etc - I have preset template which I follow to gather information and tailor my advice.I advise them of implications of pregnancy to their condition and vice versa and any risk mitigation I am able to offer them currently or prior to conception. - Medication - […] discuss meds with GP […] - Optimising medical condition […] effect o previous pregnancy or pre existing medical condition on future pregnancies and there own life - Optimising medical condition, medication advice , care during pregnancy and labour - […] addressing prescription medicine related issues […] - Pre-con clinic - full medical and obstetric history, medication review and discussion about any medications that need to be stopped/changed, family history, […] discussion about disease specific maternal and obstetric risks. In endocrine clinic - discuss pregnancy plans and then discuss contraception options or provide pre-conception advice and information about pregnancy - […] medication review if relevant - Teratgenicity of AEDs, medication, […] advised re support from ESNs thoughout pregnancy - We discuss impact of disease in pregnancy and pregnancy on disease. We discuss the impact on the type of pregnancy to be expected in terms of hospital appointments. Some discussion at this stage about the type of birth (planting a seed) common misconception need to have c-section. We also discuss medication use in pregnancy. […] - […] medication review - Yes all of the above and also discussing their specific medical condition and how pregnancy might affect it and indeed how the medical condition might impact on the pregnancy. - […] optimisation other health conditions […] - at the very broadest level at regular reviews I would discuss […] that there is a teretogenic risk with the medications and that it is ideal to have a conversation with us about 6 m before getting pregnant. Would revisit it periodically in normal reveiws and especially if they have just told me they are in a serious relationship/engaged or getting married!!) If we were having a more indepth pre-conception conversation I would discuss their understanding/prior knowledge […] depending on their medication I may do a baseline medication blood level. Advice on what to do when they find out they are pregnant. For men on valproate I would discuss the latest information. - disease and its effects on pregnancy and vice versa, medication, birth , effects on baby - […] medication […] - […] potential issues with any reg meds they already take - […] medication - […] medication , long tern medical conditions - medical condition, expectations or pregnancy, medications […] - […] medication review […] management of long-term conditions […] - […] medication safety and pregnancy risks and brief outline of care - review of medical condition and what it would mean for pregnnacy with advice on what needs to be changed prior to pregnancy and then plan for ongoing support in pregnancy […] - usually condition specific - […] drugs prescribed […] - Maternal medicine network resources. NHS resources and websites. uKTIs |
| Mental health | 10 | - […] and mental health - […] emotional well being advice - […] maternal mood and fathers mood - […] mental health […] - […] mental health […] - […] mental health factors, identifying vulnerable patients that may need additional support - […] stress […] - Also include it as part of medical history assessment ref to […] mental health […] - Mental health […] - Optimising […] mental health […] |
| Folic acid | 57 | - 5mg Folic Acid preconceptually […] - […] use of high dose folic acid for women with epilepsy […] - […] folic acid […] - […] folic acid […] - […] Folic acid […] - […] starting folic acid […] - […] Folic acid advice […] - Folic acid […] - Folic acid […] - Folic a […] - Folic acid […] - Folic acid […] - Folic acid including higher dose […] - Folic acid […] - Folic acid […] - Folic acid […] - Folic acid […] - Folic acid […] - Folic acid […] - Folic acid […] - Folic acid […] - Folic acid […] - Folic acid […] - Folic acid […] - Folic acid […] - Folic acid […] - Folic acid […] - Folic acid […] - Folic acid […] - Folic acid […] - Folic acid […] - Folic acid […] - […] includes folic acid […] - Mainly folic acid - […] folic acid […] - […] Folic acid - […] folic acid […] - […] folic acid […] - […] folic acid […] - […] FA […] - advice around folic acid […] - […] We would then discuss folic acid use […] - folic - folic acid […] - folic acid […] - folic acid […] - folic acid […] - folic acid […] - folic acid […] - folic acid […] - folic acid […] - folic acid […] - folic acid […] - preconception advice covering folic acid […] - […] folic acid […] - […] folic acid - […] folic acid […] |
| Smoking, alcohol consumption, and drug misuse | 35 | - […] advice re: smoking cessation, alcohol cessation […] - […] Smoking / alcohol […] - […] Drugs and Alcohol - […] smoking […] alcohol […] - […] smoking and alcohol […] - […] smoking and alcohol […] - […] smoking alcohol […] - […] alcohol smoking […] - […] drugs , alcohol, smoking […] - […] stopping smoking […] - […] importance of healthy lifestyle overall (stopping smoking/recreational drug use, EtOH intake […] - […] smoking, alcohol […] - […] smoking, alcohol […] - […] smoking […] - […] ETOH, smoking, recreational drugs […] - […] smoking, alcohol […] - […] smoking […] alcohol, drugs […] - […] smoking, alcohol […] - […] smoking- alcohol-recreational drugs […] - […] smoking […] - Smoking cessation […] - Smoking, alcohol intake […] - […] smoking […] - […] smoking and alcohol […] - […] smoking cessation, alcohol reduction/ceessation when trying to conceive - […] smoking and alcohol […] - […] smoking/substance use […] - […] smoking cessation and substance use […] - […] smoking, alcohol […] - […] smoking alcohol […] - […] smoking and alcohol cessation […] - […] smoking […] - smoking, drug alcohol use […] - […] smoking […] drugs prescribed and illicit - […] alcohol,smoking […] |
| Contraception | 15 | - Contraception - those appropriate for breastfeeding - […] Contraception advice and information […] - […] contraception […] - […] contraception cessation - […] contraceptive options - Optimise health for both partners for several months before removing LARC to conceive […] - […] contraception plan […] - […] discuss contraception options […] - […] contraception […] - […] contraception […] - […] I would discuss contraception […] - […] contraception […] - […] maintaining contraception until optimized health - […] contraception […] - […] contraception […] |
| BMI/weight | 31 | - BMI […] - […] weight […] - […] weight […] - […] weight […] - […] bmi […] - […] weight […] - […] weight […] - […] Healthy Weight […] - […] maintaining health weight […] - […] weight […] - […] BMI within the normal range […] - […] BMI […] - […] weight […] - […] bmi […] - […] w eight […] - […] weight […] - […] weight […] - […] weight management […] - […] weight […] - […] weight […] - […] weight […] - […] weight […] - […] discussion about general pre-pregnancy health (weight […] - […] weight optimisation […] - […] pregnancy optimisation – weight […] - […] general health advice - eg obesity - […] weight […] - […] weight […] - […] We discuss optimisation of health wight […] - Weight […] - weight […] |
| Physical activity | 9 | - […] physical activity […] - […] exercise […] - […] exercise […] - […] exercise […] - […] exercise […] - […] exercise […] - […] exercise […] - […] exercise […] - […] exercise […] |
| Diet and nutrition | 32 | - […] diet information […] Vitamin D supplementation […] - […] vitamin D […] - Advice on vitamins and diet and benefits of each - Diet/ lifestyle […] - […] vitamin D […] - […] vitamin D […] and diet - […] vitamin D […] - […] vit D […] diet […] - […] healthy eating […] supplements […] - […] healthy eating […] - […] eating well […] - […] diet […] - […] nutrition […] - […] pregnancy vitamin supplement, health diet […] - […] vit d […] - […] vit D […] - […] vitamin D […] caffeine […] - […] vitamin d […] - […] diet […] vitamin D - […] diet […] - […] Vitamin D ,Iron ,Calcium - […] Pregnancy vitamins for 8w before LARC removal and until conceive - […] vitamins […] - […] diet […] vit D […] - […] vitamin D […] - […] diet […] - […] other supplements […] - […] supplements […] - Supplements […] - […] diet […] diet and vitamins - […] diet […] - […] but D […] |
| Relationships and support | 14 | - […] support from loved ones […] - […] Relationships […] - […] partner support - […] partner support […] - […] support networks - […] taking partner support when deciding to conceive - […] Partner Health - […] partner health […] - […] Usually will briefly say same advice for male partners. - […] partner support […] - […] partner support […] - […] family support, community support - […] village of support - social network support […] |
| Immunisations | 5 | - […] vaccination schedules […] - […] Immunisations […] - […] vaccination history […] - […] rubella […] - […] vaccination status […] |
| Other | 41 | - […] antenatal care […] - […] home situation […] - Discuss this with couples participating in fertility research - […] Sometimes - alternatives to pregnancy (adoption, surrogacy) if pregnancy particularly high risk - […] offer investigations if appropriate. what a pregnancy may look like for the individual/what type of care will be provided and any extra support - […] BP check […] - […] inter pregnancy interval - […] partner smoking […] when to seek fertility support […] - […] dental health. Plus diet & exercise with male partner - […] xrays […] - […] skincare (retinols/retinoids, salicylic acid) […] - […] Physical health […] - […] General […] - Interpregnancy intervals, return of fertility - […] physical health, healthy behaviours, parenting - My support is more general than specific as they have usually birthed within the year - Optimising physical […] health […] - […] how to optimise chance of conception- - Patient led at moment, but hope to have proactive holistic approach in time. - […] social history […] - Reproductive options e.g. invasive testing, preimplantation genetic testing […] - […] breastfeeding […] - […] ideal timings post pregnancy - We discuss optimisation of health […] preparing for pregnancy - […] cervical smear, inter pregnancy interval after CS - […] discuss feeding options at antenatal contact […] - […] and impact of age on conception and pregnancy outcomes […] - […] wide-ranging - […] general "keeping yourself healthy" advice […] - Discussion - […] time in range […] - […] pregnancy following a c-section - […] personal and maternal health […] - […] physical health, […] preparation and readiness for another baby […] - […] lifestyle advice - pregnancy optimisation […] - […] domestic violence […] - […] safety advice, feeding advice - usually just one thing, whatever I can weave easily into the consultation - […] seeking GP care before pregnancy if higher risk […] - […] Sexual Health […] |

#### Table S5. Digital patient/public facing resources, N=31

| **Category** | **Count** | **Responses** |
| --- | --- | --- |
| Diabetes | 3 | - Diabetes UK have good resources […] - DiabetesUK Pregnancy, Breakthrough T1D Pregnancy, Tommy’s Pregnancy & Diabetes, DRWF Pregnancy - https://www.diabetes.org.uk/sites/default/files/2023-07/Diabetes%20UK%20Information%20Prescription%20-%20Contraception%20and%20pregnancy.pdf |
| Epilepsy | 6 | - […] Epilepsy - Different support groups depending on medical condition e.g epilepsy action - Epilepsy action - Epilepsy action pages and www.womenwithepilepsy.org.uk (I think! website by Kim Morley if you can't find it!) Epilepsy pregnancy register […] - […] Epilepsy Action \ Epilepsy Society\Young Epilepsy - We have an email that we have constructed for women with epilepsy who are pregnant, but it is also relevant to use preconception, it has links to MHRA website regarding anti-seizure medication (ASM) in pregnancy and also Epilepsy Action webpages which deal with pregnancy and medication and pre-conception. |
| Other conditions | 2 | - […] specialist medical societies as as required - Unique Rare Chromosome Organisation (https://rarechromo.org/). Cleft Lip and Palate Association (https://www.clapa.com/) |
| Tommy’s | 8 | - Tommy's […] - Tommy's App - Tommy's and "healthier together: ready for pregnancy" - Tommy's pregnancy planning - Tommy's website - Tommys app - […] Tommy's - tommys website and app - […] Tommy’s Pregnancy & Diabetes […] |
| NHS | 6 | - NHS - NHS […] - NHS Pregnancy website […] - NHS website - […] https://www.nhs.uk/contraception […] - https://www.nhsinform.scot/healthy-living/womens-health/middle-years-around-25-to-50-years/pregnancy-and-maternity/planning-for-pregnancy/ |
| RCOG and NICE | 3 | - […] if they are very keen for information I may also give the link to the RCOG guidelines - RCOG mainly […] - RCOG/NICE […] |
| Other (local) websites | 12 | - Brook website - Contraception choices - Contraceptive choices planning pregnancy page - https://www.contraceptionchoices.org/ […] - Just One Norfolk website - […] BUMPs - […] BUMPS, various Gender servcie info links - […] Squeezy app, maternity trauma support, IAPT , contraception services and individual need resources and this can include the partners especially with emotional or mental health support too. Promoting good recovery and taking this forward for future healthy pregnancy - UKTIS […] - https://www.birminghamsolihullics.org.uk/pre-conception - https://www.medicinesinpregnancy.org/ - https://www.publichealth.hscni.net/directorates/nursing-midwifery-and-allied-health-professions/midwifery/preconception-care |

#### Table S6. Other resources for healthcare professionals and practitioners used to inform preconception advice and care, N=32

| **Category** | **Count** | **Responses** |
| --- | --- | --- |
| Tommy’s | 6 | - […] Tommy’s […] - […] Tommy’s […] - Tommy's […] - Tommy's App - Tommy's, contraceptive choices - Tommys app |
| NHS | 4 | - […] NHS planning a pregnancy. - NHS -planning your pregnancy guidelines […] - […] nhs website - https://www.nhsinform.scot/healthy-living/womens-health/middle-years-around-25-to-50-years/pregnancy-and-maternity/planning-for-pregnancy/ |
| Diabetes | 4 | - Diabetes specific support websites - Planning for Pregnancy with Diabetes advice from DiabetesUK, Breakthrough T1D […] and DRWF - Study reports ie NPID - diabetes uk […] |
| Epilepsy | 4 | - Epilepsy Action - Epilepsy and Having a Baby leaflet and Women and Epilepsy. - […] Epilepsy and Pregnancy Register. MHRA epilepsy medicines in pregnancy review. MHRA Valproate and Topiramate patient guide and annual risk acknowledgement form, pregnancy prevention programme. FSRH contraception guide for drugs of teratogenic potential. - […] we also look at epilepsy action www.epilepsy.org.uk - www.womenwithepilepsy.com […] |
| Royal Colleges | 2 | - RCGP and RCOG guidelines […] - RCPsych patient information leaflets […] |
| Local resources | 4 | - Local guidelines - Local policy guideline - […] trust website […] - Resources available locally via the commissioning groups |
| Other | 20 | - Ardens template for pre-conception consultation - BUMPs - CoSRH - Consultants carry out the consultations. The lead for PPCC works with The Maternal Medicine Network - Family Nurse Partnership programme materials- facilitators, visual aids, contraceptive kits. Family planning services, GUM clinic, Common youth - Genereviews data on population carrier frequencies for rare recessive disorders - Graded model of miscarriage care - Lancet, and Scottish Miscarriage Framework - Healthier Together, Preconception toolkit (now I know about it), Healthy Start […] - Most recent evidence in lierature - […] social media - […] NB Medical/Hot Topic education resources - […] UKTIS […] - Talking with health visitors and fellow peers - […] UKTIS - UKMEC, Bumps, Maternal medicine national service specification document - Unsure - https://www.ukmec.co.uk/ - none - see above […] - […] mummysstar.org |

#### Table S7. Awareness of guidelines and policies to inform preconception advice and care, N=47

| **Category** | **Count** | **Responses** |
| --- | --- | --- |
| NICE | 31 | - Cks preconceptual advice - […] Nice guidance on preconception care - NICE - NICE - NICE - NICE […] - NICE […] - NICE CG192 - NICE CKS […] - […] NICE CKS - NICE Pre-conception Advice and Guidelines - NICE […] preconception - NICE gudelines - NICE guidelines - NICE guidelines […] - […] NICE guidance on preconception care, micronutrients - NICE […] - NICE. - Nice CKS - NICE subfertility guidance - NICE depending on the condition - […] NICE […] - there are many NICE […] guidelines on maternal and fetal medicine that I would access - […] NICE hypertension in pregnancy guideline […] - NICE (for epilepsy) […] - NICE Diabetes […] - NICE diabetes […] - NICE epilepsies NG217 and quality standards - NICE guidelines on the Epilepsies, NICE Quality standards for the Epilepsies […] - Nice Preexisiting Diabetes Guidance - […] NICE Epilepsy, NICE MS […] |
| RCOG | 13 | - […] RCOG - […] RCOG guidelines […] - […] RCOG - […] RCOG Guidance on Epilepsy […] - RCOG - RCOG - RCOG - RCOG […] - RCOG […] - […] Royal College of Obstetricians […] - […] RCOG […] - […] RCOG green top for epilepsy […] - RCOG guidelines on epilepsy and diabetes, cancer in pregnancy |
| NHS | 2 | - NHS https://www.nhs.uk/pregnancy/trying-for-a-baby/planning-your-pregnancy/ - NHS website […] |
| Other, specific to healthcare conditions | 16 | - European Society of Cardiology pregnancy guideline. European Association of the Study of the Liver pregnancy guideline. NICE hypertension in pregnancy guideline. https://www.sciencedirect.com/science/article/pii/S1569199321013382 - 2025 ESC guidelines, The ART guideance from the combined societies I was part of the authorship - Depending on medical condition […] - Depends on the medical condition like diabetes colitis haematogical conditions etc - […] specialist services ( early pregancy, fetal medicine, endocrine, cardiac etc.) - NICE (for epilepsy) […] - NICE Diabetes […] - NICE diabetes […] - NICE epilepsies NG217 and quality standards, nutritional guidance, RCOG green top for epilepsy Epilepsy Action benchmarking tool. - […] Local diabetes in pregnancy policy […] - NICE guidelines on the Epilepsies, NICE Quality standards for the Epilepsies […] - National Pregnancy in Diabetes Audit - Nice Preexisiting Diabetes Guidance - RCOG guidelines on epilepsy and diabetes, cancer in pregnancy - […] ILAE, NICE Epilepsy, NICE MS […] - sbl pid looking at pre-existing diabetes pre-con |
| Other, including Public Health Agency, audits | 19 | - DoH Sexual Health Strategy for NI. Sexual Health action plan 2023-2026 - HFEA guidance on preimplantation genetic testing - HSC PHA preconception care .Folic acid -one of life's essentials -PHA - […] We have local shared care guidelines to use with the mother and and a local policy - […] Ardens template and I'm part of the UCL IfWH SRH team - […] saving babies lives - […] FIGO, International Glucose Consensus Statement, International Confederation of Midwives Guidelines, NMC Code of Conduct - PHA guidelines - […] local guidelines - […] will also use speciality guidelines. local maternal medicine maternity network guidance - SIGN […] MHRA - Trust guidelines - UK mec, I cash - UKMEC - one key question project - safe sleep lullaby trust Healthy Child Programme Working together to Safeguard Children BFI Unicef - there are many […] national and international guidelines on maternal and fetal medicine that I would access - there are myriad across different specialist societies […] charities - […] WHO […] |

#### Table S8. Use of guidelines and policies to inform preconception advice and care, N=25

| **Category** | **Count** | **Responses** |
| --- | --- | --- |
| NICE | 9 | - NICE - NICE - NICE CG192 […] - NICE epilepsies NG217 and quality standards […] - NICE guidance - NICE guidelines […] - NICE […] - Nice diabetes and preconception - […] nice diabetes guidelines |
| RCOG | 3 | - […] RCOG […] - […] RCOG green top for epilepsy - RCOG […] |
| Other: Related to health conditions | 6 | - […] shared care guidlines (toolkit) downloadable from www.womenwithepilepsy.org.uk - NICE CG192 […] Tommy's resources on planning pregnancy with SMI - NICE epilepsies NG217 and quality standards […] RCOG green top for epilepsy - Nice diabetes […] - depends on the conditions - neg hypertension in pregnancy - diabetes in pregnancy - IBD in pregnancy - […] local diabetes in pregnancy policy - nice diabetes guidelines |
| Local resources | 3 | - Local shared care guidelines (southampton) […] - PHA -NI publications NHS - […] local diabetes in pregnancy policy […] |
| Other: Same as above (guidelines and policy they are aware of) | 17 | - All listed - All of the above - All of the guidelines previously listed - As above - I cash website for information checking - NHS https://www.nhs.uk/pregnancy/trying-for-a-baby/planning-your-pregnancy/ - […] nutritional guidance […] - […] BMC renal care guidelines - […] WHO, FIGO - National Pregnancy in Diabetes Audit guideline - ESHRE - UKMEC - as above - oKQ - same as previously listed - saving babies lives […] - there are myriad |

#### Table S9. Evaluation of preconception advice and care or services, N=18

| **Category** | **Count** | **Responses** |
| --- | --- | --- |
| Audit, survey, feedback from patients and service users | 12 | - Local audit - Local audit - As part of regular patient satisfaction surveys - Audit advice given - Audit and service evaluation - Audit carried out approx 2 yearly to check levels of advice given out. Nowhere near target of 97% - In service audit and patient feedback - Patient survey and annual report - We are currently running a pre-conception patient satisfaction survey - Audit - feedback - Friends & Family Feedback |
| As part of other programmes | 3 | - […] As part of the MMC there is basic evaluation of PPC - We have an excellent working relationship with our local sexual health team (AXCESS) they undertake the evaluation. - as part of the FNP program |
| Other | 4 | - CORE-10 outcome measure - Tick box on our online documentation to state we have discussed - We collate a data base ourselves regarding the consultations we have and then subsequent pregnancy for two services Renal and Rheumatology. […] - data collation of clients contraceptive choices and subsequent pregnancies. case study discussion/presentation. QI plans |

#### Table S10. Digital documentation of preconception advice and care, N=32

| **Category** | **Count** | **Responses** |
| --- | --- | --- |
| Letter | 9 | - As a letter - Clinic letter - Clinic letter to patient/gp - Dictate letter- stored electronically - I write a letter to the patient describing our consultation and also copy to the GP […] - LETTER FROM ENDOCRINOLOGIST - Letter to the patient documenting our discussion and my recommendations, cc'ing and GP and any other professionals. Letter gets stored in the digital notes, alongside an additional brief note about the interaction for other professionals to see. - pPC template for clinic letter that is shared with the woman and her specialist/ GP - usually paper and letter to the patient which will go to the GP |
| Electronic Health Record | 18 | - Cerner - Dictate letter- stored electronically - EMIS ,freetext within consultation programme and entry to template fro specific medication advice on avoiding pregnancy for relevant medications eg sodium valproate - EPIC - Electronic Patient Record, patient contact - Epic - Generically ,within maternal health on digital records - […] We use EPIC - In EPR - I have a personalised pre-con template - System 1 free text box, usually just write "preconception and antenatal counselling given " - SystmOne and Ardens templates. - WISDM on Welsh Clinical Portal - always good practice do document what was discussed, we use EMIS; I free text what I have said - free text, also an option contraception attendance tick box template on excelicare - on trust electronic record - system 1 - system one - Ardens template - regularly use for postnatal checks and occ for pre-conception |
| Content: Related to health conditions | 4 | - As I don't proactively provide preconceptual advice, I would document based on the presenting complaint or smi review. That might be mh, drugs, DV, medical optimisation of a condition.. - Clinical history \ Summary of discussion and action plan \ Medicine risk assessment and information \ Prescription and care recommendations - Diabetes specific impacts on the preconception period and early pregnancy when diabetes can increase the risk of adverse pregnancy outcomes. - […] specific medication advice on avoiding pregnancy for relevant medications eg sodium valproate |
| Other | 4 | - If patient attended, brief synopsis of the advice given and plan for a pregnancy. - Report discussion had with patient - We have a smart phrase that includes the information discussed at the consultation - What was discussed and what the women is currebtly feeling |

#### Table S11. Digital documentation of preconception advice and care, N=17

| **Category** | **Count** | **Responses** |
| --- | --- | --- |
| Concerns around preconception care | 9 | - I believe the pre-conception in my local trust is very minimal. One of lead clinicians runs clinics on a voluntary basis in addition to her salaried workload. - I feel PPC is extremely under recognised portion of care we provide to women and families and I feel if we get this right this has a lasting effect not just for pregnancy but for the whole lifecourse and family - In the development of the national maternal medicine service specification there is little included about funding for PCC. Even though this is a critical part of optimising care for the women […] - Thank you I feel there is a serious lack of awareness of healthy diet and lifestyle that impacts pregnancy without people even knowing. Dehydration vitamin deficiencies, poor health and lifestyle who has negative impacts that I see every day - The advice isn’t getting to women or others who have never been pregnant in a timely manner - The service we have is very much needed and has an important role to play in aiming to ensure a woman's health is optimised before pregnancy that will hopefully result in a positive outcome for mom & baby. The service is running with 2 part time midwives and we are "fighting" for more hours that will help keep time frames for appointments shorter. - Unfortunately we have little patient initiated opportunity to discuss preconceptual care as most patients move straight form non pregnant to booking into antenatal servicies with no contact with GPs. They will not have planned their pregnancies or have no idea that they could prepare to optimise their health before becoming pregnant, will have sought private fertility treatment without our knowledge or opportunity to provide such input. It is not a QOF funded activity and thus not prioritised within GMS care provision. - We need to talk more about preconception care, there is still enormous ignorance around it. - service has increased demand over last 2 years. most are women whose medical condition is well controlled and are motivated to plan pregnancy. still huge gap in reaching out to many groups who remain unprepared for pregnancy - this needs to be well before maternity services - eg primary care, paediatrics etc |
| Concerns around preconception care: Inequalities | 5 | - At the moment the bulk of my referrals come from fertility or colleagues who know me. I am concerned that it is challenging to ensure an equitable service in pre-con - I have a real interest in this and work in a region with major health inequality, so anything you come up with that might help please do feed back - […] There is also difficulties in terms of deciding what is equitable as some patients are referred from private fertility services and then being referred to the NHS clinics for PCC advice. - The ethnic minority of population may find it a challenge to access information,mainly because of language barrier. I am not aware if there are any local groups set up to provide advise to the ethnic minority e.g.in temples ,mosques. […] - There is a large amount of variation between the maternal medicine networks about how preconception care is delivered. In Y&H currently Cat C medical condition patients can be referred for specialist preconception counselling but otherwise all Cat B/A conditions it is left up to the local units. There is no funding currently where I work for preconception clinics. I think this is a big equity issue as people in some parts of the UK are being offered preconception counselling and others are not. |
| Further details on content | 2 | - […] I also assess the drugs patients are on with regards to safety and advise when and how to stop if needed, I also discuss and refer for alternatives to pregnancy eg adoption and surrogacy etc I also discuss any genetic implications and if relevant talk about PGD or cord blood sampling dependent on the case - It would be useful to have a checklist either online or paper to remind what to cover and to share with the patients/couple |
| Other | 3 | - […] I also believe that education on preconceptions should be introduced as part of school curriculum earlier on so raising awareness in the young adolescent population about preconception. - This has acted as another reminder to be more proactive in considering preconceptual care advice or optimisation, even if it's not explicitly the focus, if I can use a mental template of factors to consider covering that would help a patient in the preconceptual period. I also really hope to seek out patients to support in perhaps a targeted way like those with pre-existing mh problems etc... - With the changing of rules regarding the prescribing of Sodium Valproate for men, we are doing far more preconception counselling for men than we have done in the past |

### Supplementary File 1: online survey

**What preconception health advice, care and services are provided in the UK?**

Thank you for your interest in taking part in this study.

This study aims to map and describe the provision of preconception health advice, care and services across publicly funded or contracted health and social care settings.

We are asking health professionals and practitioners in the UK to fill out this survey to help us:

- Evaluate the extent to which current policies and guidelines are known and implemented
- Develop case studies to share best practice examples
- Identify health service gaps and develop recommendations for clinical practice, policy and research

Completing this survey takes 10-20 minutes.

Please read the Participant Information Sheet for more information and to help you decide if you would like to take part.

Please start the survey if you would like to take part.

**Who can take part?**

Health professionals and practitioners involved in providing preconception health advice and care in publicly funded or contracted health and social care settings in the UK are invited to participate in this study. This includes, but is not limited to, those working in:

- Primary and community care (e.g. general practice, community pharmacy, family hubs, children’s centres)
- Maternity, sexual and reproductive health, and health visiting services
- Secondary care and specialist services for chronic or long-term health condition management (e.g. cardiovascular disease, diabetes, mental health, epilepsy, eating disorders, endometriosis, infertility, inflammatory bowel disease, obesity)

Preconception health advice and care refers to: screening, risk assessment, education, counselling, treatment and/or advice provided to people of reproductive age – regardless of gender and pregnancy intention – with the goal of optimising their health prior to a potential pregnancy.

**Please complete the questions below to confirm your eligibility to take part:**

1. Are you a health professional or practitioner working in a publicly funded or contracted health or social care setting in the UK?

*This includes, but is not limited to, working in primary care, maternity, sexual and reproductive health, and health visiting services, and secondary care or specialist services.*

- No [Unfortunately you are not eligible to take part in this study]
- Yes

2. Do you provide preconception advice or care?

*Preconception advice and care refers to: screening, risk assessment, education, counselling, treatment, and/or advice provided to people of reproductive age – regardless of gender and pregnancy intention – with the goal of optimising their health prior to a potential pregnancy.*

- No [Unfortunately you are not eligible to take part in this study]
- Yes

* indicates questions are required / compulsory.

**Profession and clinical practice setting**

3. What is your (main) profession? *

- GP
- Practice nurse
- Health visitor
- Link worker / social prescriber
- Midwife
- Obstetrician / gynaecologist
- SRH doctor
- SRH nurse
- Community pharmacist
- Practice pharmacist
- Hospital pharmacist
- Dietician
- Psychologist
- Psychiatrist
- Substance misuse/addiction services practitioner
- Mental health practitioner
- Specialist nurse, please specify …
- Specialist doctor or consultant, please specify …
- Other, please specify: …

4. How long have you worked in this profession? *

- Less than 5 years
- 5-10 years
- 11-15 years
- 16+ years
- Prefer not to answer

5. In what setting do you (mostly) work? *

- General practice
- Community pharmacy
- Women’s Health Hub
- Family Hub
- Children’s Centre
- Sexual and reproductive health service
- Mental health service
- Substance misuse/addition service
- Health visiting
- Maternity services (early pregnancy unit)
- Maternity services (antenatal/birth/postnatal)
- Fertility clinic
- Hospital specialist service
- Other, please specify: …

**Target population and preconception advice and care content**

6. Who do you provide preconception advice and care to (target population)? (*tick all answers that apply*) *

- Adolescent girls (age 10 - 19 years)
- Adolescent boys (age 10 - 19 years)
- Adolescent non-binary or transgender people (age 10- 19 years)
- Women (age 20+ years)
- Men (age 20+ years)
- Non-binary or transgender people (age 20+ years)

7. Do you discuss preconception health with these patients/people based on pregnancy intention? (*tick all answers that apply*) *

- I discuss preconception health with all patients/people of reproductive age
- I discuss preconception health with patients/people who are intending to conceive
- Other, please specify: …

8. If relevant, please describe your target population further. For example, do they have a specific health condition such as diabetes or epilepsy? (*optional*)

Open answer:

9. How do you reach the target population? (*tick all answers that apply*) *

- Patients/people actively seek preconception advice and care
- Patients/people are referred to me through other health professionals and services
- I run a specific preconception care service/clinic
- I proactively embed preconception advice and care into relevant consultations
- Other, please specify: …

If yes to question 9 – answer option 4:

10. As part of which relevant consultations do you proactively embed preconception advice and care? (*tick all answers that apply*) *

- Medication review
- Health condition review (e.g. asthma, diabetes, thyroid, mental health)
- Contraception
- Fertility
- Sexual health
- Routine health check (e.g. as part of smoking/drinking/lifestyle)
- Cervical smear
- Postnatal check
- Other, please specify: …

11. How do you provide preconception advice and care? (*tick all answers that apply*) *

- Face-to-face consultation
- Online or phone consultation
- SMS or email
- Promotion of preconception health campaigns/information
- Other, please specify: …

12. What patient/public facing resources do you use to provide preconception advice and care? (*tick all answers that apply*) *

- None
- Signposting to website or app
- Social media
- Poster or leaflet
- Video
- Checklist
- Other, please specify: …

13. If relevant, please provide links to any digital patient/public facing resources you use. (*optional*)

Open answer:

14. How would you describe the content of the preconception advice and care you provide? For example, what topics do you discuss (e.g. folic acid, weight, medication, partner support, etc)? *

Open answer:

**Frequency of preconception advice and care provision**

15. On average, approximately how often do you provide preconception advice and care as part of your role? *

- Daily
- Weekly
- Monthly
- Less than monthly
- Other, please specify: …

16. What is the frequency of contact with a patient/person who you support as part of preconception advice and care? (*tick all answers that apply*) *

- One off consultation / conversation
- As part of ongoing care
- Other, please specify: …

17. For approximately how long have you provided preconception advice and care? *

- Less than 5 years
- 5-10 years
- 11-15 years
- 16+ years
- Don’t know

18. For approximately how long has the service you currently work for provided preconception advice and care? *

- Less than 5 years
- 5-10 years
- 11-15 years
- 16+ years
- Don’t know

19. How do you name and advertise your preconception care service? (*optional*)

Open answer:

**Clinical guideline and policy awareness, relevance and implementation**

20. Are you aware of local, national or international guidelines or policies that could inform the preconception advice and care you provide? *

- No
- Yes

If yes to question 20:

21. Please list the guidelines or policies you are aware of. (*optional*)

Open answer:

If yes to question 20:

22. Are you using any of the guidelines or policies you listed in day-to-day clinical practice to inform the preconception advice and care you provide?

- No
- Yes

If yes to question 22:

23. Please list the guidelines or policies you use. (*optional*)

Open answer:

24. Are there other resources for health professionals and practitioners that you use to inform the preconception advice and care you provide? (*optional*)

Open answer:

**Funding or commissioning model**

25. Who commissions the preconception advice and care or service you provide? (*tick all answers that apply*) *

- Not formally commissioned
- Integrated Care Board
- Local authority / local council
- NHS Board
- Don’t know
- Other, please specify: …

26. Are incentives (financial or other) in place for you to provide preconception advice and care? *

- No
- Don’t know
- Yes, please specify: …

27. If relevant, approximately when was the funding or commissioning first started? (*optional*)

Open answer:

28. Is the preconception advice and care or service you provide evaluated? *

- No
- Yes
- Don’t know

If yes to question 28:

29. Please describe how the preconception advice and care or service you provide is evaluated. (*optional*)

Open answer:

**Reporting and monitoring**

30. Are you required to document the preconception advice and care you provide as part of the clinical care process? *

- No
- Yes

If yes to question 30:

31. How do you document the preconception advice and care you provide? (*tick all answers that apply*) *

- Digital written notes
- Digital preconception care template
- Other, please specify: …

32. If you document your advice and care digitally, please provide further details, such as what you report, how, and where (e.g. the system or template you use). (*optional*)

Open answer:

33. Are you required to report the preconception advice and care you provide for audit and/or service evaluation? *

- No
- Yes

If yes to question 33:

34. Please provide further details, such as what you report, and how this is used. (*optional*)

Open answer:

**Demographic characteristics**

35. Where do you work? *

- England - London region
- England - South East region
- England - South West region
- England - West Midlands region
- England - East Midlands region
- England - East of England region
- England - North West region
- England - North East region
- England - Yorkshire and the Humber region
- Wales - Aneurin Bevan University Health Board
- Wales - Betsi Cadwaladr University Health Board
- Wales - Cardiff and Vale University Health Board
- Wales - Cwm Taf Morgannwg University Health Board
- Wales - Hywel Dda University Health Board
- Wales - Powys Teaching Health Board
- Wales - Swansea Bay University Health Board
- Scotland - NHS Ayrshire & Arran
- Scotland - NHS Borders
- Scotland - NHS Dumfries & Galloway
- Scotland - NHS Fife
- Scotland - NHS Forth Valley
- Scotland - NHS Grampian
- Scotland - NHS Greater Glasgow & Clyde
- Scotland - NHS Highland
- Scotland - NHS Lanarkshire
- Scotland - NHS Lothian
- Scotland - NHS Orkney
- Scotland - NHS Shetland
- Scotland - NHS Tayside
- Scotland - NHS Western Isles
- Northern Ireland - Belfast Health and Social Care Trust
- Northern Ireland - Northern Health and Social Care Trust
- Northern Ireland - Northern Ireland Ambulance Service HSC Trust
- Northern Ireland - South Eastern Health and Social Care Trust
- Northern Ireland - Southern Health and Social Care Trust
- Northern Ireland - Western Health and Social Care Trust
- Other, please specify: …

36. What age group are you in? *

- 18-20
- 21-30
- 31-40
- 41-50
- 51-60
- 61-70
- Over 70
- Prefer not to answer

37. How would you describe your gender? *

- Woman
- Man
- Non-binary
- Other, please specify: …
- Prefer not to answer

38. What is your ethnic background? *

- White
- Irish traveller
- Mixed/multiple ethnic groups
- Asian/Asian British
- Indian
- Pakistani
- Bangladeshi
- Chinese
- Any other Asian background
- Black/African/Caribbean/Black British
- Arab
- Any other ethnic group
- Prefer not to answer

39. Can we contact you for further details about your answers (for example to develop best practice case studies, and refine the recommendations we develop)? *

- No
- Yes

40. Can we contact you to inform you about future research studies in this area? *

- No
- Yes

If yes to question 39 and/or 40:

41. What is your name: *

Open answer:

If yes to question 39 and/or 40:

42. What is your email address: *

Open answer:

43. Would you like to share any other comments or thoughts? (*optional*)

Open answer:

### Supplementary File 2: Exploratory analyses by healthcare profession

No subgroup analyses were planned as part of this study due to the low pragmatic target sample size (n=75).

To inform future research, analyses were conducted to explore potential differences in preconception care provision by healthcare professional.

These findings should be interpreted with caution: cell counts are very low (mostly < n=5) and the study sample was not intended to be a representative sample of healthcare professionals providing preconception care across the UK.

Exploratory analysis suggest:

- Digital communication: Online and phone communication was highest among Diabetes Specialist Nurses (100%) and GPs (91.7%), with SMS/email communications highest among GPs (50%). Among professionals who reported online and phone communication and SMS/email communication, they were lowest among SRH doctors (45.5%) and specialist epilepsy nurses (14.3%), respectively.
- Population: In general, SRH doctors (54.5 %) and specialist nurses (specialist epilepsy nurses: 42.9%) reported greater inclusion of adolescent non-binary or transgender people (10-19) than primary care groups (GPs: 33.3%).
- Preconception care was mainly triggered by pregnancy intention for pharmacists (100%), SRH doctors (90.9%) and GPs (83.3%), whereas specialist nurses (85.7–100%) and health visitors (80%) more often delivered routine, population-based care.
- Frequency: Weekly preconception care was most commonly provided by SRH doctors (63.3%), while daily preconception care was most common among health visitors (40%) and specialist epilepsy nurses (33.3%). Frequency of advice and care reported by GPs varied widely.

Preliminary conclusion based on these findings: specialist services provide more routine and inclusive care, while primary care is more opportunistic and intention-led, with greater use of digital follow-up.

Data availability: <https://doi.org/10.5258/SOTON/D3968>.
